# Monitoring multi-pathogens and SARS-CoV-2 variants in aircraft and airport wastewater

**DOI:** 10.1101/2024.05.11.24307221

**Authors:** Martin Tay, Benjamin Lee, Muhammad Hafiz Ismail, Jerald Yam, Dzulkhairul Maliki, Karina Yew-Hoong Gin, Sae-Rom Chae, Zheng Jie Marc Ho, Yee Leong Teoh, Lee Ching Ng, Judith Chui Ching Wong

## Abstract

**Background:** As global travel resumed in COVID-19 endemicity, the potential of aircraft wastewater monitoring to provide early warning of disease trends for SARS-CoV-2 variants and other infectious diseases, particularly at international air travel hubs, was recognized. We therefore assessed and compared the feasibility of testing wastewater from inbound aircraft and airport terminals for 18 pathogens including SARS-CoV-2 in Singapore, a popular travel hub in Asia.

**Methods:** Wastewater samples collected from inbound medium- and long-haul flights and airport terminals were tested for SARS-CoV-2. Next Generation Sequencing (NGS) was carried out on positive samples to identify SARS-CoV-2 variants. Airport and aircraft samples were further tested for 17 other pathogens through quantitative reverse transcription polymerase chain reaction (RT-qPCR).

**Results:** The proportion of SARS-CoV-2-positive samples and the average virus load was higher for wastewater samples from aircraft as compared to airport terminals. Cross-correlation analyses indicated that viral load trends from airport wastewater led local COVID-19 case trends by two to five days. A total of ten variants (44 sub-lineages) were successfully identified from aircraft wastewater and airport terminals, and four variants of interest (VOIs) and one variant under monitoring (VUM) were detected in aircraft and airport wastewater 18-31 days prior to detection in local clinical cases. The detection of five respiratory and four enteric viruses in aircraft wastewater samples further underscores the potential to expand aircraft wastewater to monitoring pathogens beyond SARS-CoV-2.

**Conclusion:** Our findings demonstrate the feasibility of aircraft wastewater testing for monitoring infectious diseases threats, potentially detecting signals before clinical cases are reported. The triangulation of similar datapoints from aircraft wastewater of international travel nodes could therefore serve as a useful early warning system for global health threats.

## Introduction

Globalisation, urbanisation and population mobility can accelerate the spread of infectious diseases in a highly-connected world. This has been demonstrated in the COVID-19 pandemic where air travel facilitated the rapid and extensive transmission of SARS-CoV-2 globally [1]. Although restrictions on international travel [2] and mandated clinical tests at airports for incoming travelers [3] had some success in minimising new introductions of the virus [4–6] and bought many countries or states time for implementation of vaccination, such controls had adverse impacts on the economy of states and were thus not sustainable. Following the reopening of borders, countries across the globe have experienced successive waves of COVID-19 transmission, largely driven by the continued emergence of new SARS-CoV-2 variants such as Delta and Omicron BA.5, among others [7], which are more transmissible with high rates of immune breakthrough. [8]. These waves of transmission continue to pose threats of increased transmissibility, immune breakthrough [9] and varying disease severity [10, 11], bringing uncertainty to society [12], economies [13], and health care systems [14]. Additionally, respiratory viruses and infectious diseases like respiratory syncytial virus (RSV), tuberculosis and influenza that have been previously contained due to early pandemic measures are now re-emerging [15, 16]. This therefore underscores the need for comprehensive and timely public health surveillance of circulating COVID-19 variants and other pathogens, to guide the calibration of response measures and inform policy decisions [14].

Wastewater surveillance is an emerging tool for public health surveillance and has been used to complement clinical surveillance in the monitoring of infectious diseases trends [17–19]. It has been used to track the circulation of SARS-CoV-2, as well as other respiratory [20], gastrointestinal [21], and other infectious diseases [22, 23]. It facilitates early case identification [24], situational assessment [17], and monitoring of public health trends of various infectious diseases in a population [25]. While wastewater monitoring has been largely focused on testing samples collected from wastewater treatment plants and manholes to understand the prevalence of infections in local communities, expanding the approach to aircraft wastewater holds significant potential. The testing of flights from high-traffic destinations and strategic travel nodes could provide an early indication of global transmission trends and serve as an early alert system for emerging or re-emerging infections [1, 26–28].

Several countries including Australia [29], the United States (US) [30], the United Kingdom [31], France, Denmark [32] and the United Arab Emirates [33] have conducted SARS-CoV-2 testing on aircraft wastewater. Among them, the US, France, and Denmark have further conducted sequencing to identify variants from the aircrafts, serving as an indication of the circulating variants originating from the inbound cities. Although the findings were focused on SARS-CoV-2 monitoring, the studies collectively affirm the feasibility of the approach for monitoring emerging infectious diseases threats, and experts advocate for the establishment of a global wastewater surveillance consortium to promote alignment of methodologies and best practices to facilitate monitoring and data sharing across regions [34].

In this study, we conducted aircraft and airport wastewater surveillance at Singapore Changi airport which serves as a popular mall destination for locals [35] and a travel node in Asia [36]. We monitored 17 other pathogens beyond SARS-CoV-2 and its variants from January 2023 to February 2023. In this pilot study, we detected nine other pathogens in aircraft wastewater collected from medium- and long-haul flights, demonstrating the utility of aircraft wastewater surveillance for monitoring respiratory and gastrointestinal diseases, in addition to the monitoring of SARS-CoV-2 variants.

## Methods

### Wastewater sampling from lavatory service truck

Surveillance of inbound flights were conducted from January 2023 to February 2023, categorised into 26 medium-haul flights (flight duration ≤ 8 h) sampled for the first month and 15 long-haul flights (flight duration > 8 h) sampled for the second month (Table 1), originating from two separate continents respectively. Categorisation of flight duration was guided by previous studies [37, 38]. A lavatory service truck was designated for the collection of samples from aircraft. From the lavatory service truck, samples were transferred into 250 mL screw-capped bottles using a portable peristaltic pump (Masterflex, Germany) and transported to the laboratory for testing. The collection tank of the lavatory service truck was disinfected and flushed between collections. A total of 19 and 15 samples were collected from medium-haul and long-haul flights, respectively. As only one lavatory service truck was deployed for this study, on occasions where multiple flights were scheduled for collection on a single day, the samples from these flights were pooled. Among wastewater samples collected from medium-haul flights, six samples comprised pooled samples from two to three medium-haul flights each.

**Table 1.** Proportion of positive SARS-CoV-2 samples and variants identified in wastewater collected from aircrafts and airport terminal sites.

| Collection Date (2023) | Site/ Flight | No. of Samples | SARS-CoV-2 Detected | Met Sequencing Analysis Requirements<sup>1</sup> | Variants Identified<sup>2</sup> |
| --- | --- | --- | --- | --- | --- |
| January | Medium-Haul | 19<sup>3</sup> | 13/19 (68%) | 1/19 (5.26%) | BA.5.2.48 |
|  | Airport (Site 1) | 36 | 8/36 (22%) | 0/36 (0%) | NA |
|  | Airport (Site 2) | 36 | 10/36 (27%) | 1/36 (2.8%) | BA.2.3.20 |
| February | Long-Haul | 15 | 9/15 (60%) | 3/15 (20%) | BQ.1.1<br>XBB.1.5 |
|  | Airport (Site 1) | 32 | 4/32 (13%) | 1/32 (3.13%) | XBB.2.4<br>XBB.1.5.X<br>XBB.1.11.X |
|  | Airport (Site 2) | 32 | 13/32 (41%) | 4/32 (12.5%) | XBB.1.5.X<br>XBB.1.9.X<br>XBB.1.11.X<br>XBB.1.18.X<br>XBB.1.22.X<br>XBL |

<sup>1</sup> Only samples with $\geq 80\%$ genome coverage, 10 times read depth are analysed
<sup>2</sup> Detailed sub-lineages are provided in Supplementary Material Table S1
<sup>3</sup> Six samples comprised pooled samples from two to three medium-haul flights

### Wastewater sampling from manholes serving the airport terminal sites

Two autosamplers were deployed at two sites in Singapore Changi Airport to draw wastewater samples from the sewage manholes covering the terminals which served both the long- and medium-haul flights (Site 1: Terminals 1 and 2; Site 2: Terminal 3). Autosamplers were programmed to collect four composite samples twice a week, with each composite sample constituting sewage drawn from the sampling point every 15 min over a 6 h period. Samples from each autosampler were transferred into 250 mL screw-capped bottles and transported to the laboratory. A total of 136 (68 each) composite samples across two airport terminal sites were collected.

### Sample processing and nucleic acid extraction

Aircraft wastewater samples were centrifuged at 10,000 *g* for 20 min and the supernatant was filtered through a 0.22 µm filter (Corning, Tewksbury, MA, USA). Wastewater samples collected from the airport terminal sites were subjected to a single centrifugation step at 4,000 *g* for 20 min. Virus concentration of the supernatant (∼15 mL) was carried out using ultrafiltration and RNA extraction as previously described [24, 39]. A duplex one-step quantitative reverse transcription polymerase chain reaction (RT-qPCR), targeting both SARS-CoV-2 nucleocapsid N1-gene and CPQ_056, was used to screen wastewater samples for the presence of SARS-CoV-2 and rule out PCR inhibitors respectively. Primer and probe sequences were as previously reported [40, 41]. PCR reactions were carried out in a final reaction volume of 20 µL containing 0.5 µM of N1 primers and 0.25 µM of N1 probe, 0.25 µM of CPQ_056 primers and 0.125 µM of CPQ_056 probe, 1X Luna® Universal Probe One-Step RT-qPCR Kit (New England Biolabs, Ipswich, MA, USA), 1X Luna® RT Enzyme Mix and 2.5 µL of template. Thermal cycling was performed on the QuantStudio 5 machine (Applied Biosystems, USA) as described in the Supplementary Material.

### Multiple-pathogen detection using panel assays and quantitative polymerase chain reaction

Wastewater samples collected from 12 aircrafts, each originating from unique airports (medium-haul, n=7; long-haul, n=5), between 8 January to 23 February 2023; and wastewater samples collected weekly from the 2 airport terminal sites (n=12) within the same period, were tested using the BioFire Respiratory Panel (RP2.1) assay as per manufacturer’s instructions (BioFire Diagnostics LLC, Saltlake, UT). Briefly, sample buffer was added to 300 µL of the ultrafiltration retentate. The sample-buffer mixture was injected into a test pouch and loaded into the BioFire Filmarray Torch Instrument (BioFire Diagnostics LLC, Saltlake, UT) for sample processing, nucleic acid extraction, amplification, and molecular detection. RT-qPCR/qPCR analysis was also performed on RNA extracts in accordance with methods described in Goh *et al.* for the detection of Aichivirus, Adenovirus and Hepatitis A virus [42].

### Next Generation Sequencing and bioinformatic analyses of SARS-CoV-2 variants

RNA extracts from wastewater samples with a minimum of 20 SARS-CoV-2 RNA copies/µL were further subjected to Next Generation Sequencing (NGS). First strand cDNA synthesis was performed followed by amplicon generation with the ARTIC v4.1 primers panel. The amplicon generation performed is an adapted version of a previously published protocol [43], and the adaptations are further described in the Supplementary Material.

The data was processed and mapped against the reference sequence (MN908947.3) using the nfcore/viralrecon pipeline [44]. The aligned bam file was then parsed through Freyja [45] to deconvolute the diversity of SARS-CoV-2 in each sample. Only samples that had a minimum of 80% genome coverage at a mapped read depth of 10X were used. SARS-CoV-2 variant information of wastewater collected from water reclamation plants and clinical cases were obtained from Singapore’s National Environment Agency and Ministry of Health, respectively.

### Correlation of COVID-19 viral load in wastewater and clinical cases

Correlation analysis of COVID-19 viral load in wastewater and clinical cases was carried out for the period of August 2022 to October 2023. The number of COVID-19 clinical cases reported globally was extracted from the complete Our World in Data COVID-19 dataset (https://ourworldindata.org/covid-cases). The LOESS (locally estimated scatterplot smoothing) method was applied to SARS-CoV-2 virus concentration detected in the community and airport terminal sites to summarise the time series observations into a smoothed curve. The SARS-CoV-2 virus concentration in community and airport terminal sites samples were obtained as described in a previous study [17]. The association of viral load in wastewater with clinical cases was assessed with the Pearson correlation coefficient between the respective wastewater datasets and the global cases or local cases. To test the effectiveness of wastewater surveillance as a leading indicator of resurgence of transmission, cross-correlation was performed between the daily clinical cases in Singapore and the smoothened curves of the wastewater datasets.

## Results

### SARS-CoV-2 variants of concern

SARS-CoV-2 RNA was detected in wastewater samples collected from both aircraft and airport terminal sites. Five variants of interest (VOI) were first detected in aircraft wastewater collected from long-haul flights (XBB.1.5.2, XBB.1.5.7, XBB.1.5.11, XBB.1.5.18, XBB.1.5.21), of which four were eventually detected in the local water reclamation plants samples between 18 to 31 days later (XBB.1.5.2, XBB.1.5.7, XBB.1.5.11, XBB.1.5.18) (**Error! Reference source not found.**). Among these VOIs, imported clinical cases for three sub-lineages (XBB.1.5.7, XBB.1.5.18, XBB.5.21) were reported 8 to 40 days after it was detected in aircraft wastewater (**Error! Reference source not found.**). Additionally, the XBB.4 variant under monitoring (VUM) was first detected in wastewater collected from the airport terminal (Site 1) prior to detection in water reclamation plants 4 days later, with the first imported clinical case reported 19 days later.

**Figure 1.**
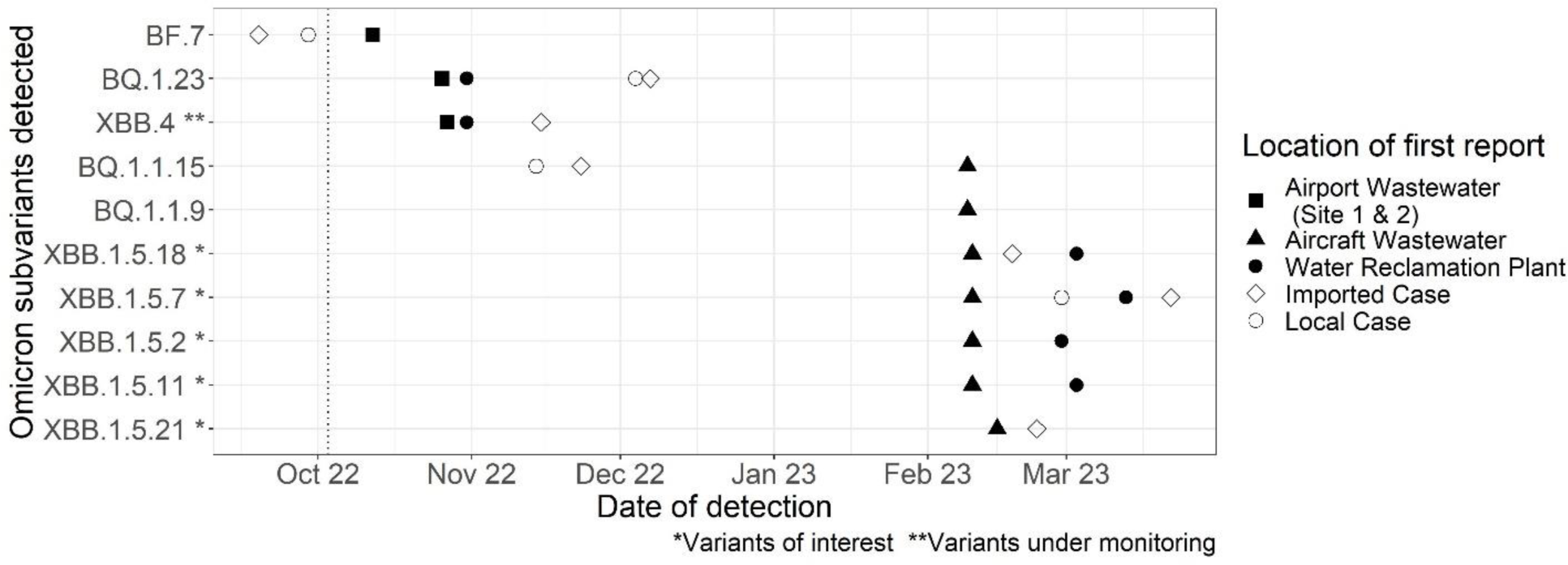
Detection of Omicron subvariants in wastewater samples collected from aircraft, airport terminal sites, water reclamation plants, and in clinical samples of reported cases. Detection of most SARS-CoV-2 variants in wastewater samples collected from aircraft (closed triangles) and airport terminal sites (closed squares), preceded the detection in wastewater samples collected from inland water reclamation plants (closed circles), and in clinical samples (imported cases: open diamond; local cases: open circles).

A total of ten variants (44 sub-lineages) were successfully identified from NGS sequencing of wastewater samples collected from airport terminal sites (40 sub-lineages) and medium- and long-haul aircrafts (4 sub-lineages) (Table 1, Supplementary Material Table S1). All the variants identified were sub-lineages of the Omicron variant and several of these sub-lineages have been classified as VOIs and VUMs by the World Health Organization (WHO). The proportion of positive SARS-CoV-2 samples in aircraft wastewater was higher (60-68%) than in wastewater collected from airport terminal sites (13-41%) (Table 1).

### Positive correlation of SARS-CoV-2 virus load from airport terminal sites with local COVID-19 cases

Comparison of wastewater viral load trends at both the airport terminal sites revealed higher positive correlation with local COVID-19 cases (*r* = 0.73 and 0.67) than those reported globally (negative correlation, *r*=-0.0047, not significant, and -0.38) (Supplementary Material Table S2). Positive correlation was also observed between the local wastewater viral load trend in the community and that of both airport terminal sites (*r* = 0.55 and 0.71) (Supplementary Material Table S3). Expectedly, the trend of local COVID-19 cases was more strongly associated with the wastewater viral load trend from autosamplers deployed in the community compared to those from airport terminal sites (*r* = 0.93 vs *r* = 0.73 and 0.67) (Supplementary Material Table S2) [17]. Nevertheless, it was interesting to note that cross-correlation analyses suggested that the wastewater viral load trends at the airport terminal sites led the trend in local COVID-19 cases by two and five days, respectively, while the wastewater viral load trend in the community led by a day (Figure C).

**Figure 2.**
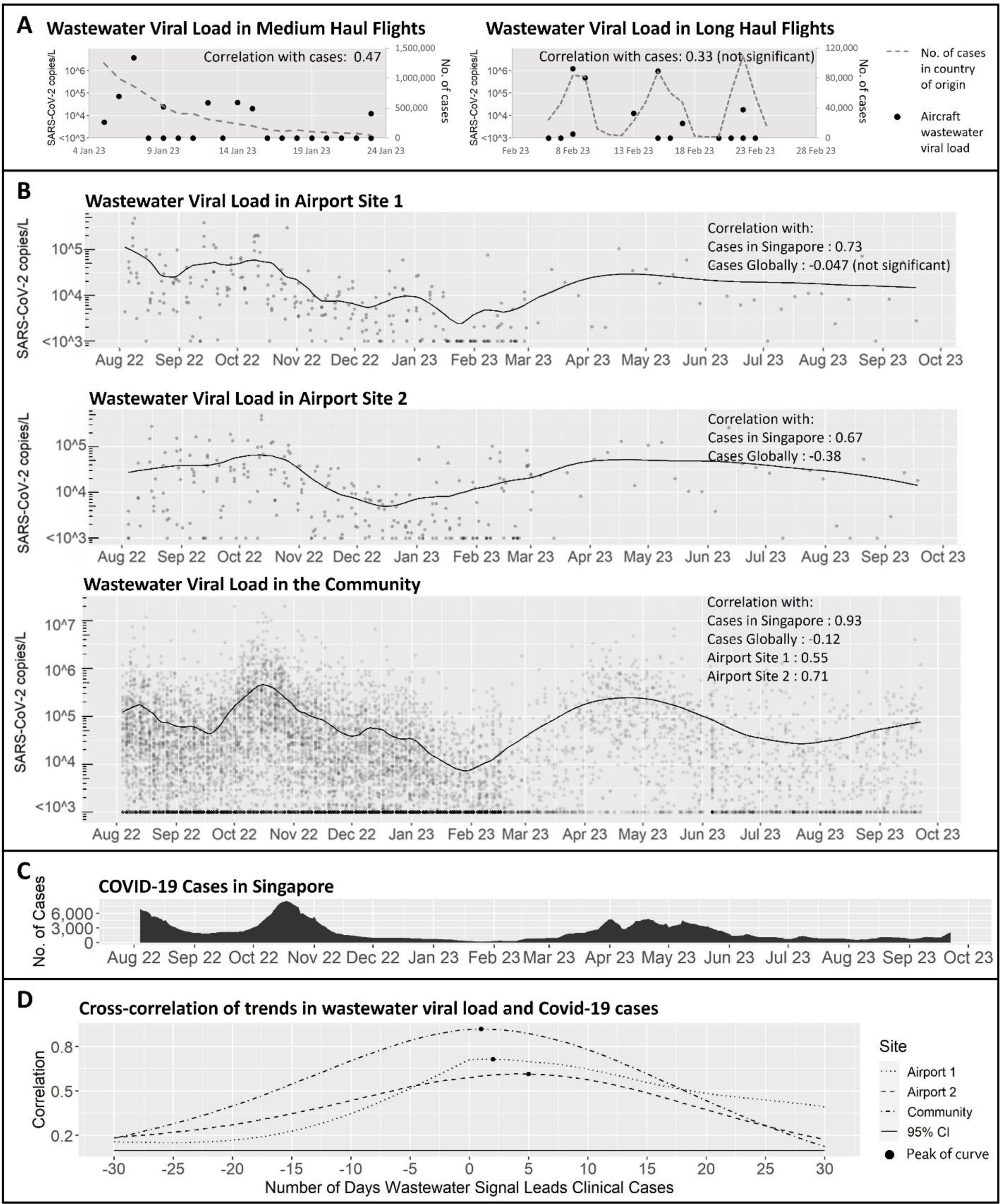
SARS-CoV-2 virus load and COVID-19 cases. Trend of SARS-CoV-2 viral load in (A) aircraft wastewater samples from medium- and long-haul flights, (B) Airport terminal sites and the community, and (C) COVID-19 cases reported in Singapore. (D) Cross-correlation of trends in wastewater viral load (airport terminal sites and community) and COVID-19 cases.

### Detection of multiple pathogens in aircraft and airport wastewater samples

All aircraft wastewater samples screened for multiple pathogens yielded positive detections of more than one pathogen. A total of five respiratory viruses (coronavirus NL63, rhinovirus/enterovirus, influenza A virus, Parainfluenza type 3 virus, Respiratory Syncytial Virus) and four enteric viruses (Norovirus, Aichivirus, Adenovirus, Hepatitis A) were detected, with enteric viruses detected at higher proportions when compared with respiratory viruses (14-86% vs 14-60%) (Figure 3).

**Figure 3.**
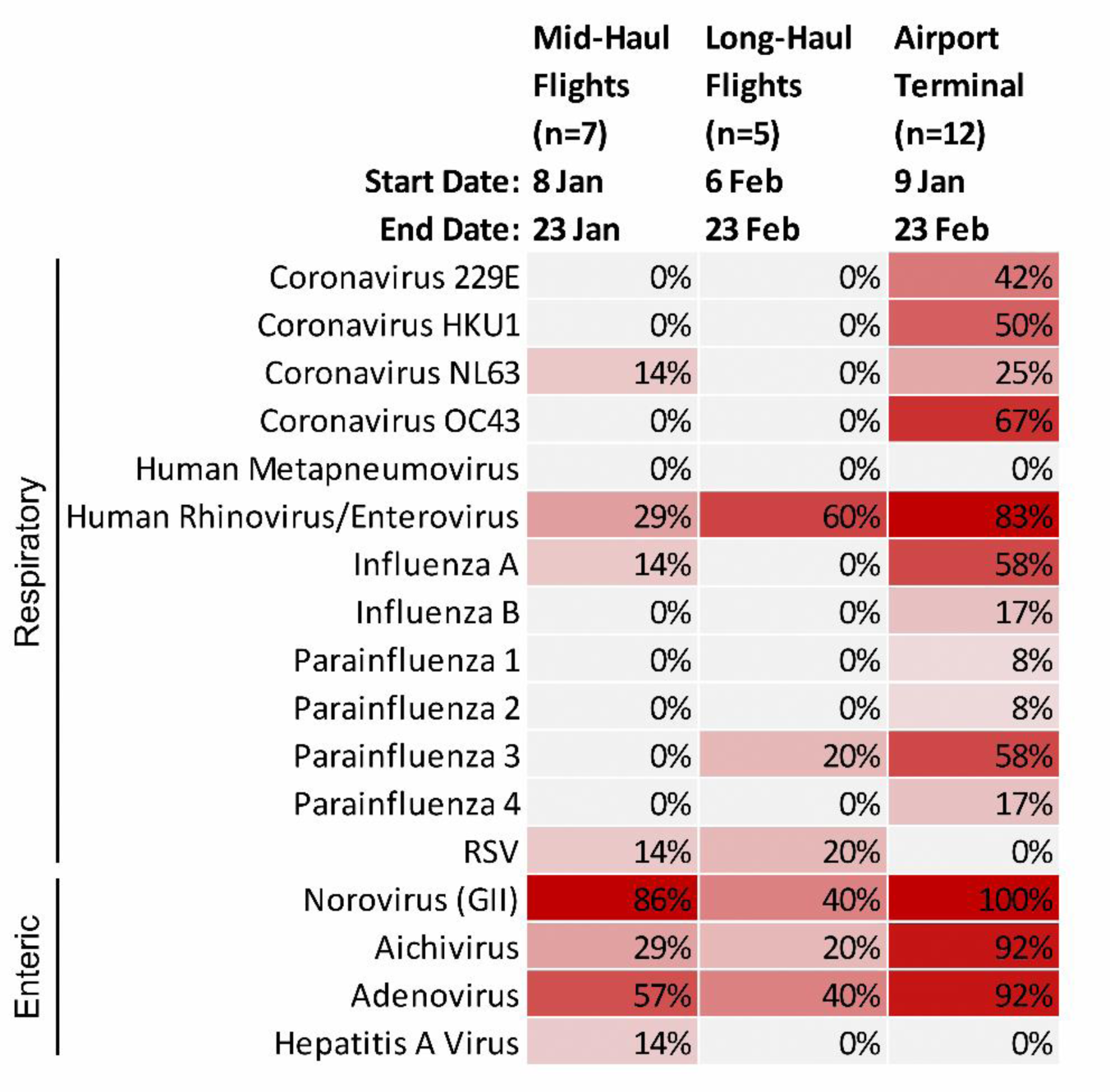
Multi-pathogens detected in aircraft and airport terminal sites wastewater samples. Samples collected from the airport terminal sites had a higher proportion testing positive and yielded more pathogens than aircraft wastewater samples. Both airport terminal sites and aircraft wastewater samples yielded higher proportions of samples testing positive for enteric viruses than for respiratory viruses.

Samples collected from the airport terminal sites had a higher proportion testing positive, and yielded more pathogens (8-100%, n=14), when compared with aircraft wastewater samples (mid-haul flights, 14-86%, n=8; long-haul flights, 20-60%, n=6). Both airport terminal sites and aircraft wastewater samples had higher proportion of samples tested positive for enteric viruses (14-100%) than for respiratory viruses (8-83%) (Figure 3).

## Discussion

The detection of some SARS-CoV-2 variants in aircraft wastewater preceded the detection in wastewater samples collected from inland water reclamation plants and in clinical samples. Specifically, the detection of various VOIs (i.e. sub-lineages of XBB.1.5.X) in long-haul flights preceded the detections in inland wastewater and clinical samples by 31 days and 18 days, respectively. Furthermore, a VUM (XBB.4) was also detected in airport terminal sites’ wastewater 4 and 19 days earlier than detections in inland wastewater and clinical samples, respectively. Collectively, the findings suggest the usefulness of aircraft and airport wastewater surveillance to potentially provide early signals of new virus introductions. This approach would be particularly useful for monitoring more transmissible or deadly variants, especially if surveillance was carried out at frequencies which could provide insights on likely introductions to the country.

Sampling wastewater from airport terminals have been suggested as a proxy for monitoring aircraft wastewater, particularly if there were capacity or resource constraints [46]. However, our study revealed that the viral load trends from airport terminal sites’ wastewater samples correlated more strongly with the trends in community cases rather than global cases; potentially providing an indication of the trend of COVID-19 infections in the community instead. Cross-correlation analyses of viral load trends from airport terminal sites and community wastewater samples showed that wastewater signals were observed to be lead indicators of local COVID-19 case trends, further highlighting the utility of wastewater surveillance as an important supplemental to clinical surveillance systems. Notably, the detection of more pathogens in wastewater from airport terminal sites compared to aircraft could possibly be due to the combined wastewater coverage from travellers from multiple regions as well as the local community. In our settings, we were limited by the availability of suitable sampling points in the airport, and the sampling points could not segregate sewage lines serving the aircrafts from those serving the airport. As the airport in Singapore comprises retail outlets that are popular with the local community [35], and are staffed mostly by resident workers, these could partially explain the corroboration of trends in wastewater viral load and community cases. While the testing of our airport terminal sites’ wastewater samples may provide limited insights on the global situation, its utility could be increased at airports with segregated sampling points and with lower traffic from the community.

The successful detection of respiratory and enteric viruses in aircraft wastewater samples underscores the utility of aircraft wastewater monitoring for other infectious diseases beyond SARS-CoV-2. The monitoring of medium- and long-haul flights also revealed unique trends of the respective regions. For example, norovirus and adenovirus were found at a higher proportion in medium-haul flights originating from the same continent, as compared to rhinoviruses/ enteroviruses which were detected at a higher proportion in long-haul flights. Overall, these trends could reflect the epidemiology and circulating viruses of the originating regions. When medium- and long-haul flights were further compared, we found that the proportion of positive SARS-CoV-2 samples (44% vs 60%) and the sequencing rate (1% vs 8%) were lower for medium-haul flights. One possible explanation could be the lower likelihood of toilet visits, and in turn, virus shedding, for passengers onboard shorter flights [37].

This study has a few limitations. The scope of the study was a pilot study limited to surveillance of 19 medium- and 15 long-haul flights from January to February 2023. The toilet use patterns of the travelers were unknown and it was not possible to infer virus shedding rates in wastewater, and in turn, disease incidence. The study was also limited by the availability of sampling trucks and the design of the sewer network at the airport. On the latter point, it was thus not possible to segregate wastewater collected from travelers from that of the local community.

Despite the potential of aircraft wastewater monitoring, our study has revealed challenges and the need for more efficient sampling and testing solutions. Sampling has been largely carried out via lavatory service trucks or grab sampling [31–33], with only a few companies offering devices capable of sampling directly from the aircraft with minimal disruption to the short turnaround time (90-120 min for large aircraft and 25-40 min for smaller aircraft) [29, 30, 47, 48]. Aircraft wastewater is less diluted than regular sewage and presents challenges in sample preparation [29]; while the wastewater sample may have a higher viral or bacterial load, the concentrated wastewater may lead to clogged virus concentration filters and inhibition from disinfectants, among others. Variant analyses via NGS also have limited sensitivity in detecting SARS-CoV-2 in samples with low viral loads, especially in complex matrices such as aircraft wastewater. Mutations within viral genomes may also affect the binding efficiency of amplification primers, potentially leading to low coverage rates across the genome. The accuracy of deconvolution tools, like Freyja used in this study, is also dependent on existing lineage information obtained by prior clinical sequencing data and published sequences [45]. Lastly, the complicated process of NGS sample preparation prior to its subsequent analyses, and the turn-around-time required for sequencing may be a bottleneck, delaying the availability of results and subsequent public health responses.

While conducting wastewater testing at airport terminals and aircrafts may offer an early warning of diseases for the local community, modelling analyses reveal that data from a network of airports are needed to provide early warning and situational awareness of emerging outbreaks [49]. It is therefore needful to encourage innovation in wastewater sampling and testing technologies, and to work as a network in a global initiative. This collaboration would yield synergistic effect, enhancing global surveillance and situational awareness of emerging pathogens.

## Author Contribution

MT, BL, MHI: data curation, formal analysis, investigation, methodology, writing – original draft. JY, DM, KYHG: formal analysis, investigation. SRC, ZJMH, TYL: conceptualization. NLC: Supervision, conceptualization, methodology. JWCC: Supervision, conceptualization, methodology, formal analysis, writing – original draft. All authors provided critical review and revision of the manuscript and were responsible for the decision to submit for publication.

## Data Availability

The data underlying this article will be shared on request and may be subject to approval and/or a data sharing contract.

## Acknowledgements

The authors thank the Wastewater-Based Epidemiology Branch and Molecular Biotechnology Branch of the Microbiology and Molecular Epidemiology Division in the National Environment Agency for their support in wastewater sampling and testing of SARS-CoV-2 and other pathogens. The authors appreciate Mr. Alvin Xie Cheng Goh for his assistance with variant monitoring of clinical samples. The authors also thank the Changi Airport Group and SIA Engineering company for facilitating the collection of aircraft wastewater samples. The authors appreciate the Public Utilities Board, Singapore’s Water Agency, for their provision of water reclamation plant samples and their support in access to the sewer network.

## References

1. Sokadjo, Y.M. and M.N. Atchadé, The influence of passenger air traffic on the spread of COVID-19 in the world. Transportation Research Interdisciplinary Perspectives, 2020. 8: p. 100213.

2. Bielecki, M., et al., Air travel and COVID-19 prevention in the pandemic and peri-pandemic period: A narrative review. Travel Medicine and Infectious Disease, 2021. 39: p. 101915.

3. Yokota, I., P.Y. Shane, and T. Teshima, Logistic advantage of two-step screening strategy for SARS-CoV-2 at airport quarantine. Travel Medicine and Infectious Disease, 2021. 43: p. 102127.

4. Le, T.-M., et al., Framework for assessing and easing global COVID-19 travel restrictions. Scientific Reports, 2022. 12(1): p. 6985.

5. Wilson, N., et al., Estimating the impact of control measures to prevent outbreaks of COVID-19 associated with air travel into a COVID-19-free country. Sci Rep, 2021. 11(1): p. 10766.

6. Ge, Y., et al., Impacts of worldwide individual non-pharmaceutical interventions on COVID-19 transmission across waves and space. International Journal of Applied Earth Observation and Geoinformation, 2022. 106: p. 102649.

7. Beesley, L.J., et al., SARS-CoV-2 variant transition dynamics are associated with vaccination rates, number of co-circulating variants, and convalescent immunity. eBioMedicine, 2023. 91.

8. Samieefar, N., et al., Delta Variant: The New Challenge of COVID-19 Pandemic, an Overview of Epidemiological, Clinical, and Immune Characteristics. Acta Biomed, 2022. 93(1): p. e2022179.

9. McLean, G., et al., The Impact of Evolving SARS-CoV-2 Mutations and Variants on COVID-19 Vaccines. mBio, 2022. 13(2): p. e0297921.

10. Yang, B., et al., Comparison of control and transmission of COVID-19 across epidemic waves in Hong Kong: an observational study. Lancet Reg Health West Pac, 2024. 43: p. 100969.

11. Abdool Karim, S.S. and T. de Oliveira, New SARS-CoV-2 Variants — Clinical, Public Health, and Vaccine Implications. New England Journal of Medicine, 2021. 384(19): p. 1866–1868.

12. UnitedNations. COVID-19 SOCIO-ECONOMIC IMPACT. 2023 [cited 2023; Available from: https://www.undp.org/coronavirus/socio-economic-impact-covid-19.

13. Wei, X., L. Li, and F. Zhang, The impact of the COVID-19 pandemic on socio-economic and sustainability. Environ Sci Pollut Res Int, 2021. 28(48): p. 68251–68260.

14. OECD, Ready for the Next Crisis? Investing in Health System Resilience. 2023.

15. The Lancet Respiratory, M., Patterns of respiratory infections after COVID-19. The Lancet Respiratory Medicine.

16. Marani, M., et al., Intensity and frequency of extreme novel epidemics. Proc Natl Acad Sci U S A, 2021. 118(35).

17. Wong, Y.H.M., et al., Positive association of SARS-CoV-2 RNA concentrations in wastewater and reported COVID-19 cases in Singapore – A study across three populations. Science of The Total Environment, 2023. 902: p. 166446.

18. Chua, F.J.D., et al., Co-incidence of BA.1 and BA.2 at the start of Singapore’s Omicron wave revealed by Community and University Campus wastewater surveillance. Sci Total Environ, 2023. 875: p. 162611.

19. Mohapatra, S., et al., Wastewater surveillance of SARS-CoV-2 and chemical markers in campus dormitories in an evolving COVID - 19 pandemic. J Hazard Mater, 2023. 446: p. 130690.

20. Boehm, A.B., et al., Wastewater concentrations of human influenza, metapneumovirus, parainfluenza, respiratory syncytial virus, rhinovirus, and seasonal coronavirus nucleic-acids during the COVID-19 pandemic: a surveillance study. The Lancet Microbe, 2023. 4(5): p. e340–e348.

21. Chacón, L., et al., Wastewater-Based Epidemiology of Enteric Viruses and Surveillance of Acute Gastrointestinal Illness Outbreaks in a Resource-Limited Region. Am J Trop Med Hyg, 2021. 105(4): p. 1004–1012.

22. Wolfe, M.K., et al., Use of Wastewater for Mpox Outbreak Surveillance in California. N Engl J Med, 2023. 388(6): p. 570–572.

23. Wong, J.C.C., et al., Zika Surveillance Complemented with Wastewater and Mosquito Testing. Social Science Research Network [Preprint] Available at SSRN: https://ssrn.com/abstract=4512915 or 10.2139/ssrn.4512915, 2023.

24. Wong, J.C.C., et al., Non-intrusive wastewater surveillance for monitoring of a residential building for COVID-19 cases. The Science of the total environment, 2021. 786: p. 147419–147419.

25. Diamond, M.B., et al., Wastewater surveillance of pathogens can inform public health responses. Nature Medicine, 2022. 28(10): p. 1992–1995.

26. Bogoch, I.I., et al., Anticipating the international spread of Zika virus from Brazil. The Lancet, 2016. 387(10016): p. 335–336.

27. Hosseini, P., et al., Predictive Power of Air Travel and Socio-Economic Data for Early Pandemic Spread. PLOS ONE, 2010. 5(9): p. e12763.

28. Tian, H., et al., Increasing airline travel may facilitate co-circulation of multiple dengue virus serotypes in Asia. PLOS Neglected Tropical Diseases, 2017. 11(8): p. e0005694.

29. Ahmed, W., et al., Wastewater surveillance demonstrates high predictive value for COVID-19 infection on board repatriation flights to Australia. Environ Int, 2022. 158: p. 106938.

30. Morfino, R.C., et al., Notes from the Field: Aircraft Wastewater Surveillance for Early Detection of SARS-CoV-2 Variants - John F. Kennedy International Airport, New York City, August-September 2022. MMWR Morb Mortal Wkly Rep, 2023. 72(8): p. 210–211.

31. Farkas, K., et al., Wastewater-based monitoring of SARS-CoV-2 at UK airports and its potential role in international public health surveillance. PLOS Glob Public Health, 2023. 3(1): p. e0001346.

32. Qvesel, A.G., et al., *SARS-CoV-2 Variants BQ.1 and XBB.1.5 in Wastewater of Aircraft Flying from China to Denmark*, *2023*. Emerg Infect Dis, 2023. 29(12): p. 2559–2561.

33. Albastaki, A., et al., First confirmed detection of SARS-COV-2 in untreated municipal and aircraft wastewater in Dubai, UAE: The use of wastewater based epidemiology as an early warning tool to monitor the prevalence of COVID-19. Sci Total Environ, 2021. 760: p. 143350.

34. Murakami, M., et al., The growing need to establish a global wastewater surveillance consortium for future pandemic preparedness. J Travel Med, 2023. 30(7).

35. Hamdi, R. Singapore’s Changi Is Changing the Idea of What an Airport Can Be. 2019 [cited 2024; Available from: https://skift.com/2019/03/11/singapores-changi-is-changing-the-idea-of-what-an-airport-can-be/.

36. Cripps, K. Singapore’s Changi Airport fully reopens Terminal 2 following dramatic makeover. 2023 [cited 2024; Available from: https://edition.cnn.com/travel/singapore-changi-airport-terminal-2-expansion/index.html.

37. Jones, D.L., et al., Suitability of aircraft wastewater for pathogen detection and public health surveillance. Science of The Total Environment, 2023. 856: p. 159162.

38. Wilkerson, J.T., et al., Analysis of emission data from global commercial aviation: 2004 and 2006. Atmos. Chem. Phys., 2010. 10(13): p. 6391–6408.

39. Mailepessov, D., et al., Development of an efficient wastewater testing protocol for high-throughput country-wide SARS-CoV-2 monitoring. Sci Total Environ, 2022. 826: p. 154024.

40. Niu, C., et al., Interlaboratory assessment of quantification of SARS-CoV-2 RNA by reverse transcription digital PCR. Analytical and Bioanalytical Chemistry, 2021. 413: p. 7195–7204.

41. Stachler, E., et al., Quantitative CrAssphage PCR assays for human fecal pollution measurement. Environmental science & technology, 2017. 51(16): p. 9146–9154.

42. Goh, S.G., et al., Occurrence of microbial indicators, pathogenic bacteria and viruses in tropical surface waters subject to contrasting land use. Water Res, 2019. 150: p. 200–215.

43. D, D.N.A., et al., COVID-19 ARTIC v3 Illumina library construction and sequencing protocol v5. 2020.

44. Patel, H., et al., nf-core/viralrecon: nf-core/viralrecon v2.6.0 - Rhodium Raccoon. 2023, Zenodo.

45. Karthikeyan, S., et al., Wastewater sequencing reveals early cryptic SARS-CoV-2 variant transmission. Nature, 2022. 609(7925): p. 101–108.

46. Deere, D., et al., Ad-hoc guidance: Wastewater sampling of aircrafts for SARS-CoV-2 surveillance: A guidance document for Member States. 2023.

47. Wegrzyn, R.D., et al., Early Detection of Severe Acute Respiratory Syndrome Coronavirus 2 Variants Using Traveler-based Genomic Surveillance at 4 US Airports, September 2021-January 2022. Clin Infect Dis, 2023. 76(3): p. e540–e543.

48. Wignall, A. How it works: the aircraft turnaround. 2022 [cited 2024; Available from: https://www.aerotime.aero/articles/32767-how-it-works-the-aircraft-turnaround.

49. Jin, S., et al., Feasibility of wastewater-based detection of emergent pandemics through a global network of airports. PLOS Global Public Health, 2024. 4(3): p. e0003010.

