## Supplemental Material for "Monitoring multi-pathogens and SARS-CoV-2 variants in aircraft and airport wastewater"

### Supplementary Material

#### RT-qPCR thermocycling conditions

The thermocycling conditions are reverse-transcription at 55 °C for 10 min, PCR activation at 95 °C for 1 min, followed by 45 amplification cycles of 95 °C for 10 s and 55 °C for 30 s.

#### Next Generation Sequencing library preparation and sequencing

Extracted RNA were converted to ssDNA using the Maxima H Minus First Strand cDNA Synthesis Kit (ThermoFisher Scientific, USA) as per manufacturer's instruction, followed by amplification with the ARTIC v4.1 primers (IDT, USA). The PCR uses 2 reactions for each sample, with each reaction consisting of 12.5 µL of Phusion Flash 2X PCR Master Mix (ThermoFisher Scientific, USA), 3.6 µL of primer pool, 2.9 µL of nuclease-free water, and 6 µL of cDNA. The thermocycling conditions were 1 cycle of 98 °C for 30 s, 35 cycles of 95 °C for 15 s and 63 °C for 5 min, and held at 20 °C. RT-PCR products from the ARTIC v4.1 primer pools A and B were combined to a total volume of 50 µL and cleaned up using the AMPure XP beads (Beckman Coulter, USA) as per manufacturer's instruction. The DNA was eluted in 29 µL of nuclease-free water and 2 μL was quantified using the QuantiFluor dsDNA System (Promega, USA) on the Quantus Fluorometer (Promega,USA).

Sequencing libraries were prepared with the Nextera XT DNA Library Prep Kit (Illumina, USA) as dual-indexed paired end libraries. The libraries were quality checked and quantified using the TapeStation D1000 ScreenTape (Agilent, USA) before normalization and loaded onto the MiSeq Desktop Sequencer (Illumina, USA) as 2 x 300 bp PE libraries. The subsequent FASTQ files generated were also demultiplexed and adapter trimmed on the MiSeq prior to analysis.

**Supplementary Table S1. SARS-CoV-2 sub-lineages identified in wastewater collected from aircrafts and airport terminal sites**

| Sample Type | Sample Origin | Date | Lineage | Subvariant |
|---|---|---|---|---|
| Airport | Site 2 | 2/1/2023 | BA.2 | BA.2.3.20.12 |
| | Site 1 | 8/2/2023 | XBB | XBB.2.4 |
| | | | | XBB.1.5.14 |
| | | | | XBB.1.5.11 |
| | | | | XBB.1.11.1 |
| | Site 2 | 9/2/2023 | XBB | XBB.1.9.1 |
| | Site 2 | 13/2/2023 | XBL | XBL |
| | | | XBB | XBB.1.18.1.1 (FE.1) |
| | | | | XBB.1.9.1 |
| | | | | XBB.1.18.1 |
| | | | | XBB.1.5.24 |
| | | | | XBB.1.22.1 |
| | | | | XBB.1.22 |
| | | | | XBB.1.11.1 |
| | Site 2 | 27/2/2023 | XBB | XBB.1.5.80 |
| | | | | XBB.1.5.11 |
| | | | | XBB.1.5.92 |
| | | | | XBB.1.5.52 |
| | | | | XBB.1.5.12 |
| | | | | XBB.1.5.30 |
| | | | | XBB.1.5.2 |
| | | | | XBB.1.5.75 |
| | | | | XBB.1.5.7 |
| | | | | XBB.1.5.36 |
| | | | | XBB.1.5.13.2 (EK.2) |
| | | | | XBB.1.11.1.4 (FP.4) |
| | | | | XBB.1.5.81 |
| | | | | XBB.1.5.34 |
| | | | | XBB.1.5.62 |
| | Site 2 | 28/2/2023 | XBB | XBB.1.5.23 |
| | | | | XBB.1.5.31 |
| | | | | XBB.1.5.15 |
| | | | | XBB.1.5.65 |
| | | | | XBB.1.5.56 |

| | | | | |
|---|---|---|---|---|
| | | | | XBB.1.5.17 |
| | | | | XBB.1.5.100 |
| | | | | XBB.1.5.95 |
| | | | | XBB.1.5.96 |
| | | | | XBB.1.5.51 |
| | | | | XBB.1.5.76 |
| | | | | XBB.1.5.75 |
| | | | | XBB.1.5.12 |
| | | | | XBB.1.5.30 |
| | | | | XBB.1.5.13.3 (EK.3) |
| | | | | XBB.1.5.36 |
| | | | | XBB.1.5.84 |
| | | | | XBB.1.5.67 |
| | | | | XBB.1.5.57 |
| | | | | XBB.1.5.29 |
| | | | | XBB.1.5.14.1 (EL.1 ) |
| Medium-haul Flights | Not applicable (NA) | 7/1/2023 | BA.5 | BA.5.2.48 |
| Long-haul Flights | NA | 8/2/2023 | BA.5 | BQ.1.1 |
| | NA | 9/2/2023 | XBB | XBB.1.5 |
| | NA | 15/2/2023 | XBB | XBB.1.6 |

**Supplementary Table S2. Correlation between wastewater viral load of various sources and number of cases from different regions**

| Source of Wastewater Viral Load | Number of cases | Pearson correlation coefficient, r | p value | 95% Confidence interval |
|---|---|---|---|---|
| Medium haul flights | Country of origin (Medium haul flights) | 0.47 | 0.043 | [0.018, 0.76] |
| Long haul flights | Country of origin (Long haul flights) | 0.33 | 0.32 | [-0.33, 0.78] |
| Airport Terminal Site 1 | Singapore | 0.73 | 1.17E-68 | [0.68, 0.77] |
| Airport Terminal Site 1 | Global | -0.047 | 0.34 | [-0.14, 0.05] |
| Airport Terminal Site 2 | Singapore | 0.67 | 9.92E-55 | [0.61, 0.72] |
| Airport Terminal Site 2 | Global | -0.38 | 1.39E-14 | [-0.45, -0.28] |
| Community (Singapore) | Singapore | 0.93 | 1.93E-174 | [0.91, 0.94] |
| Community (Singapore) | Global | -0.12 | 0.014 | [-0.22, -0.025] |

**Supplementary Table S3. Correlation between wastewater viral load of the community and airport terminal sites**

| Source of Wastewater Viral Load | Pearson correlation coefficient, r | p value | 95% Confidence interval |
|---|---|---|---|
| Community (Singapore) vs Airport Terminal Site 1 | 0.55 | 6.65E-34 | [0.48, 0.61] |
| Community (Singapore) vs Airport Terminal Site 2 | 0.71 | 2.60E-65 | [0.66, 0.76] |
